# Psychopharmacolipidomics: Understanding the Global Impact of Psychopharmacologic Treatments on Plasma Lipidomic Profiles

**DOI:** 10.64898/2026.01.06.26343451

**Authors:** Annika Weber, Sergi Papiol, Anna Tkachev, Fanny Senner, Daniela Reich-Erkelenz, Alba Navarro-Flores, Mojtaba Oraki Kohshour, Urs Heilbronner, Maria Heilbronner, Monika Budde, Kristina Adorjan, Richard Musil, Jens Wiltfang, Carsten Spitzer, Max Schmauß, Eva Z. Reininghaus, Georg Juckel, Andreas Fallgatter, Udo Dannlowski, Peter Falkai, Philipp Khaitovich, Thomas G. Schulze, Eva C. Schulte

## Abstract

**Objective:** This study aimed to investigate the naturalistic effects of psychopharmacologic treatments on the plasma lipidome using a transdiagnostic approach, focusing on antipsychotics and antidepressants.

**Methods:** Using liquid chromatography-mass spectrometry, we analyzed untargeted plasma lipidomic profiles of 622 individuals (192 control individuals, 187 with schizophrenia, 243 with bipolar disorder) who participated in the naturalistic PsyCourse Study, a longitudinal deep-phenotyping study focussing on affective and non-affective psychoses recruited across Germany and Austria. Differential lipid species, class enrichments, and clustering patterns were examined using tools like match.it, lipidr and clustering methods in R to investigate associations between the plasma lipidome and different psychopharmacologic treatments.

**Results:** The lipid class enrichment analysis revealed that antipsychotics induced the most pronounced lipid alterations, affecting six classes including triacylglycerides (TAG), acylcarnitines (CAR), and diacylglycerols (DAG). TAGs were increased, while fatty acids (FA) were consistently decreased with both antipsychotic and antidepressant treatment. Although antipsychotics, antidepressants, and tranquilizers showed overall class effects, subgroups displayed differential, sometimes opposing patterns. The “ziprasidone-like” group showed increased sphingomyelins (dSM) and decreased FA. Antidepressants primarily affected TAG and phosphatidylcholines (PC), with SSRI and SNRI subgroups showing significant enrichment.

Unbiased hierarchical clustering based on sex, age, BMI, diagnosis, and the use of individual drugs revealed a cluster of severely affected individuals characterized by key lipid species (dSM 39:2, FA 26:4, CAR 14:0).

**Conclusion:** Psychopharmacologic treatments, particularly antipsychotics and antidepressants, significantly alter plasma lipid profiles, with specific lipid classes like TAGs and FAs affected consistently across medication groups but also class- and subgroup-specific effects, yielding insights into potential mechanisms of action, drug response, and unwanted effects.

## Introduction

The second most common molecules in the human brain are lipids. They are involved in a plethora of integral processes in regulating neuronal structure and function. These include but are not limited to determining membrane fluidity and permeability, vesicle formation, transport and release and cell plasticity (Lauwers et al., 2016). Brain lipid composition is also known to change throughout the human lifespan (Yu et al., 2020). Accordingly, it is difficult to imagine that lipids do not also play central roles in brain diseases like severe mental health disorders and the mechanisms of psychopharmacologic treatments used to treat them. The current understanding of the role of lipids—both so-called “clinical” (e.g. high- and low-density lipoproteins or triglycerides) and “non-clinical” (i.e. the rest of the human lipidome)—in mental health disorders, however, is still relatively limited. Earlier studies have been small in size and results have often been heterogeneous (reviewed in Tkachev et al., 2023). Studies of increasing size have recently been able to identify alterations in peripheral lipid profiles in individuals with severe mental health disorders spanning the psychotic spectrum (Cao et al., 2019; Tkachev et al., 2023; Brunkhorst-Kanaan et al., 2019). Potential lipidomics-based diagnostic panels for severe mental health disorders have also been described (Tkachev et al., 2023; Liu et al., 2021) and plasma lipidomic changes are beginning to be linked to symptom dimensions (Oraki Khoshour et al., 2025). In prior work in a large-scale lipidomics dataset of 1,552 individuals, which also included individuals from the PsyCourse Study described in this project, we identified a plasma lipid profile of 77 lipid species that transethnically differentiated individuals with schizophrenia (SCZ) from controls with high accuracy (Tkachev et al., 2023). Transdiagnostic lipidomic alterations have also been observed across disorders and distinguish affected individuals from controls with high accuracy (Tao et al., 2021; Tkachev et al., 2023). The reasons for this intriguing finding remain to be elucidated, with possible explanations ranging from shared genetics to shared life-style factors.

High on the list of possible environmental influences shaping mental-health-disorder-related plasma lipidomic profiles are medication effects. Especially changes in “clinical” lipids resulting in dyslipidemias and linked to metabolic syndromes in individuals with schizophrenia are well documented (Ijaz et al., 2018; Penninx et al., 2018), although the causal mechanisms are also not yet clear (Franz et al., 2024). Yet, analyzing the lipidomic effects of these drugs is crucial for advancing knowledge in the field (Correia et al., 2021) and specific lipid profiles have already been suggested as potential biomarkers for treatment response (Tkachev et al., 2021; Kaddurah-Daouk et al., 2007; Almeida et al., 2020; Aquino et al., 2018).

The currently available plasma lipidomics studies in psychotic disorders have accounted for medication effects to variable degrees. In the transdiagnostic, transethnic plasma lipidomic profile we identified previously, correlation was reasonably high and statistically significant in individuals with a multi-year duration of illness and continuous pharmacologic treatment compared with either first episode psychosis patients (r=0.86) or those with a longer duration of illness but without medication for more than 6 months (r=0.75). Free fatty acids (FAs), triacylglyerides (TAGs), and carnitines (CAR) differed most between these groups (Tkachev et al., 2023). Longitudinal studies at treatment initiation have shown both drug-specific (e.g., TAGs, FAs) and shared (e.g., increased PEs) effects of antipsychotics such as olanzapine, risperidone, or aripiprazole (Kaddurah-Daouk et al., 2007), and lipid signatures distinguishing treatment responders from non-responders have been identified (Kaddurah-Daouk et al., 2007; Almeida et al., 2020). Comparison of results across studies, however, is hampered by the fact that different antipsychotics were assessed. Overall, study sizes are small with mostly 50 individuals or less per group. Studies focusing on antidepressant-dependent plasma lipidomic changes in humans are even more scarce. In one of the largest studies to date, levels of several phophatidylcholines (PCs) were increased by treatment with and response to a selective serotonin reuptake inhibitor (SSRI) in 136 participants with major depressive disorder (MDD) (MahmoudianDehkordi et al., 2021).

In the work depicted herein, we perform a comprehensive analysis of the effects of psychopharmacologic treatments on plasma lipidomic profiles, focusing on three different classes of drugs—antipsychotics, antidepressants, and tranquilizers. The ultimate aim of this study is to decipher differential plasma lipidomic effects of psychopharmacologic treatment in 622 individuals of the naturalistic PsyCourse Study, providing an important component in understanding the intricate relationship between lipidomic alterations, pharmacologic treatment and affective and non-affective psychoses. The analyses depicted herein were not performed as part of Tkachev et al. (Tkachev et al., 2023) as granular data on pharmacologic treatments were only available in the PsyCourse Study but neither of the other two studies.

## Methods

### Study participants

A total of 622 individuals were selected from the full cohort (1786 individuals) of the naturalistic, deep-phenotyping PsyCourse Study (www.psycourse.de) (Budde et al., 2019), recruited across Germany and Austria, based on the availability of plasma lipidomic data. For baseline characteristics see table 1. Plasma samples for lipidomic analyses were collected throughout the day and were non-fasting. Study participants received pharmacologic treatments prescribed by their treating psychiatrists as usual with medication data recorded as specific drugs and drug categories; dosages were not considered.

**Table 1:** Baseline characteristics of the individuals included in the analysis

|  | **Bipolar disorder**<br>**(n=243)** | **Schizophrenia**<br>**(n=187)** | **Controls**<br>**(n=192)** |
| --- | --- | --- | --- |
| **Age** [years], mean (sd) | 44.9 (13.4) | 39.3 (12.8) | 38.3 (16.1) |
| **BMI** [kg/m<sup>2</sup>], mean (sd) | 28.3 (6.2) | 28.4 (6.3) | 24.3 (4.6) |
| **Female**, % (n) | 46.5 (113) | 29.9 (56) | 56.8 (109) |
| **Male**, % (n) | 53.5 (130) | 70.1 (131) | 43.2 (83) |
| **Medication use** |  |  |  |
| **Antipsychotics**, % (n) | 64.2 (156) | 92.5 (173) | 0 (0) |
| Aripiprazole, % (n) | 15.6 (38) | 25.1 (47) | 0 (0) |
| Clozapine, % (n) | 0.4 (1) | 19.3 (36) | 0 (0) |
| Haloperidol, % (n) | 1.2 (3) | 7.5 (14) | 0 (0) |
| Olanzapine, % (n) | 9.5 (23) | 13.9 (26) | 0 (0) |
| Quetiapine, % (n) | 35.0 (85) | 23.5 (44) | 0 (0) |
| Risperidone, % (n) | 2.5 (6) | 18.7 (35) | 0 (0) |
| Ziprasidone, % (n) | 5.3 (13) | 4.3 (8) | 0 (0) |
| **Antidepressants, % (n)** | **47.7 (116)** | **26.2 (49)** | **0 (0)** |
| SNRI, n (%) | 16.9 (41) | 7.5 (14) | 0 (0) |
| Venlafaxine, % (n) | 8.2 (20) | 7.0 (13) | 0 (0) |
| SSRI, n (%) | 17.7 (43) | 12.3 (23) | 0 (0) |
| Escitalopram, % (n) | 5.3 (13) | 1.6 (3) | 0 (0) |
| Sertraline, % (n) | 8.2 (20) | 8.0 (15) | 0 (0) |
| Tricyclic antidepressants, n (%) | 3.7 (9) | 2.7 (5) | 0 (0) |
| Mirtazapine, % (n) | 5.3 (13) | 3.7 (7) | 0 (0) |
| Bupropion, % (n) | 7.8 (19) | 1.1 (2) | 0 (0) |
| **Mood stabilizers, % (n)** | **69.5 (169)** | **6.4 (12)** | **2.1 (4)** |
| Lamotrigine, n (%) | 11.9 (29) | 1.1 (2) | 1.0 (2) |
| Lithium, n (%) | 31.7 (77) | 2.1 (4) | 0 (0) |
| Valproic acid, n (%) | 26.3 (64) | 3.7 (7) | 0 (0) |
| **Tranquilizers, % (n)** | **9.9 (24)** | **19.8 (37)** | **1.0 (2)** |

sd=standard deviation, SSRI=selective serotonin reuptake inhibitor, SNRI=Serotonin noradrenalin reuptake inhibitor

Written informed consent was obtained from each participant. Demographic and medication information for all participants is outlined in Table 1. The study was approved by the University Hospital Munich’s ethical committee (Application number: 17-13) as well as by those at the other study sites and was carried out in accordance with the Declaration of Helsinki.

### Lipid quantification

Plasma sample collection and lipid quantification were carried out as reported previously (Tkachev et al., 2021, Supplementary material: Lipid quantification process). 1361 lipid features were reproducibly detected and putatively annotated based on mass and retention time using an in-house library. This yielded 394 annotated lipid species (Supplementary Table S1) belonging to 16 different lipid classes: triacylglycerides (TAG), acylcarnitines (CAR), phosphatidylcholines (PC), phosphatidylcholine plasmalogens (PC-P), ceramides (dCer), phosphatidylethanolamines (PE), phosphatidylethanolamine plasmalogens (PE_P), fatty acids (FA), sphingomyelins (dSM), plasmanylphosphatidylcholines (PC-O), cholesteryl esters (CE), diacylglycerols (DAG), lysophosphatidylcholines (LPC), lysophosphatidylcholine plasmalogens (LPC_P), lysoplasmanylphosphatidylcholines (LPC_O), and lysophosphatidylethanolamines (LPE). PC-O, PE-P, and PC-P were not analyzed as separate classes in lipidr but summarized to PC and PE.

### Statistical Analyses

Lipid species not meeting normality criteria (|skewness| ≤ 1, |kurtosis| ≤ 5) were excluded (n = 27). Lipids previously reported as affected by fasting (Tkachev et al., PMID 34064997) were considered, and three species (two FA, one LPE) present in our dataset were removed (Supplementary Table S1). The remaining 364 lipids were log2-transformed, centered, and scaled using the scale function in R 4.2.1 (https://www.R-project.org/). Outliers (> ±3 standard deviations) were removed prior to analysis.

For analyses of individual lipid species, matched sample pairs were generated using the match.it package (Ho et al., 2011). Matching method depended on sample size (n < 300: nearest neighbor; n ≥ 300: optimal full matching). For medication analyses, diagnosis was always a matching criterion, with additional covariates such as age, sex, and BMI considered; healthy controls were included in some analyses. Whenever possible, patients taking a specific medication were matched to patients with the same diagnosis who were not receiving the medication. Matching success was evaluated via pairwise standard mean differences and overall propensity scores (Supplementary Table S12). Associations were assessed using t-tests and linear regression, with FDR correction (Benjamini-Hochberg).

To evaluate robustness, a second analysis was conducted at the lipid-class level. Matched groups for medication classes (antipsychotics, antidepressants, tranquilizers), diagnosis (SCZ, BPD), and individual drugs (≥15 patients per single-drug group) were analyzed using lipidR 2.15.1 (Mohamed et al., 2020). These analyses included differential analysis of annotated lipids and lipid-class enrichment, serving as a validation of species-level findings.

To account for multiple drug use, hierarchical clustering was performed using a Gower distance matrix and the hclust function in R, based on sex, age, BMI, diagnosis, and use of individual drugs (valproic acid, lamotrigine, haloperidol, clozapine, olanzapine, risperidone, quetiapine, aripiprazole, ziprasidone, venlafaxine, sertraline, mirtazapine, bupropion, escitalopram, lithium). A four-cluster solution was selected based on elbow and silhouette plots (Supplementary Figures F1–F2). Covariate distributions (sex, age, BMI, diagnosis, medication use, and clinical measures: CGI, PANSS, GAF, YMRS, IDS-C) were evaluated across clusters using ANOVA for numeric and chi-square tests for categorical variables. Associations between lipid species and clusters were assessed via ANCOVA, followed by a 10,000-permutation test for lipid-class enrichment. Finally, a random forest model was applied after removing highly correlated lipids (r > 0.9), with feature importance used to identify key cluster-associated lipid species.

## Results

### Plasma lipidomic effects of drug classes

Covariate balance was generally good across groups, with minor residuals in antipsychotics for sex/diagnosis (Supplementary Table S12). Lipid class enrichment showed antipsychotics with the broadest effects: 6 classes altered (↑DAG/dCer, ↓CAR/FA/PC, TAG heterogen; n=329 matched). Antidepressants affected 4 classes (↑dCer/TAG, ↓LPC/PC; n=165), tranquilizers 3 (↑TAG, ↓CAR/FA; n=63); mood stabilizers showed no changes (n=185) and are therefore not shown in Figure 1A and were not carried forward in this work for this reason. Shared patterns included TAG↑ across classes except stabilizers, dCer↑ in antipsychotics/antidepressants, FA↓ in both, and CAR↑ in antipsychotics/tranquilizers (Figure 1A)

**Figure 1.**
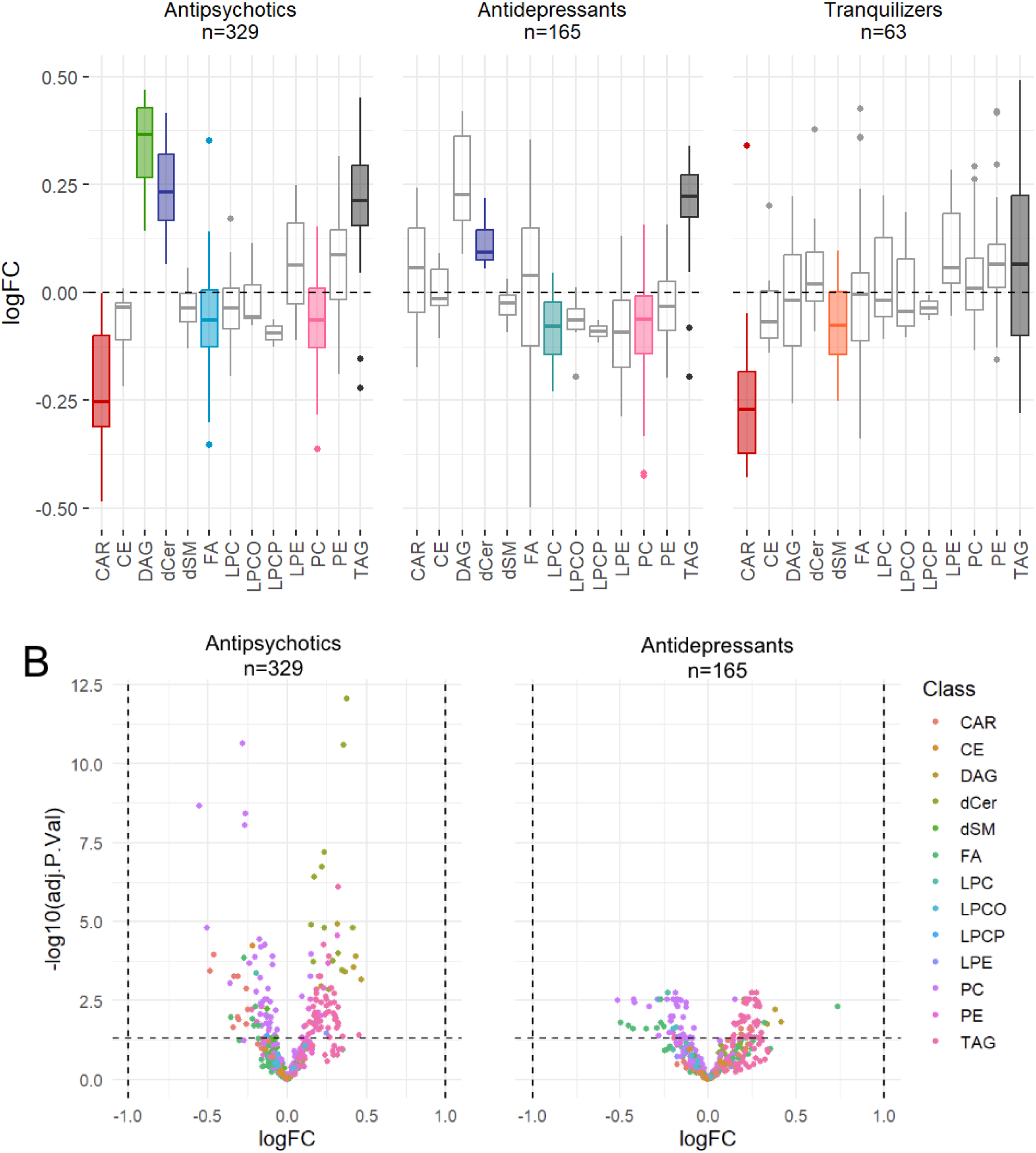
Lipid class alterations associated with psychotropic medication use **Lipid class enrichment analysis** in matched samples of individuals taking antipsychotics, antidepressants, mood stabilizers, or tranquilizers. Colored boxplots represent lipid classes with significant enrichment (Benjamini–Hochberg corrected *p* < 0.05). Matching was performed using full matching as implemented in the MatchIt package. *N* = total number of individuals included in each analysis. logFC=log fold change, CAR=acylcarnitines, CE=cholesteryl esters, DAG=diacylglycerol, dCer=ceramides, dSM=sphingomyelins, FA=fatty acids, LPC=lysophosphatidylcholines, LPCO= lysoplasmanylphosphatidylcholines, LPCP=lysophosphatidylcholine plasmalogens, LPE= lysophosphatidylethanolamines, PC=phosphatidylcholines, PE=phosphatidylethanolamines, TAG=triacylglycerides.

Differential analysis confirmed antipsychotics’ dominance (153 significant species vs. 98 antidepressants, 0 tranquilizers; Figure 1B, Table 2, Supplementary Table S7), spanning CARs (e.g. CAR 10:2), DAGs (e.g. DAG 34:1, 34:3, 36:1, 36:3, 36:5), dCers (e.g. dCer 32:1, 34:2, 38:1, 42:2, 42:4, 43:2, 43:3, 44:2), dSMs (e.g. dSM 39:1, 41:2), FAs (e.g. FA 10:2), LPCs, LPEs, PCs, PCOs, PCPs, PEs, PEPs, and TAGs (baseline characteristics: Supplementary Tables S2–S4).

**Table 2:** 10 Lipid species with the lowest adjusted p-values in the differential analysis for the antipsychotics group (left) and antidepressants group (right)

| Antipsychotics |  |  | Antidepressants |  |  |
| --- | --- | --- | --- | --- | --- |
| Lipid species | Lipid class | Adjusted p-value | Lipid species | Lipid class | Adjusted p-value |
| dCer 36:2 | dCer | 8,73E-13 | LPC 18:2 | LPC | 0,00176 |
| PCP 34:2 | PC | 2,36E-11 | PCO 36:2 | PC | 0,00176 |
| dCer 36:1 | dCer | 2,52E-11 | TAG 55:6 | TAG | 0,00176 |
| PCP 35:2 | PC | 2,19E-09 | TAG 56:5 | TAG | 0,00176 |
| PCP 36:2 | PC | 3,91E-09 | LPE 18:2 | LPE | 0,00293 |
| PCP 34:1 | PC | 9,12E-09 | LPE 18:1 | LPE | 0,00293 |
| dCer 40:2 | dCer | 6,51E-08 | dCer 36:1 | dCer | 0,00293 |
| dCer 42:3 | dCer | 1,82E-07 | PC 40:4 | PC | 0,00293 |
| dCer 34:1 | dCer | 3,83E-07 | PCO 42:7 | PC | 0,00293 |
| TAG 56:5 | TAG | 7,96E-07 | TAG 50:3 | TAG | 0,00293 |

### Plasma lipidomic effects of drugs with similar mechanisms of action

Antipsychotics were grouped by receptor profiles: “ziprasidone-like” (ziprasidone, risperidone, paliperidone) and “clozapine-like” (clozapine, olanzapine); antidepressants into SSRIs (fluoxetine, sertraline, escitalopram, paroxetine, citalopram), SNRIs (milnacipran, duloxetine, reboxetine, venlafaxine), TCAs (amitriptyline, doxepine, clomipramine, tianeptine, trimipramine), NaSSAs (mirtazapine) (baseline characteristics: Supplementary Tables S4–S5). Matching was based on utilization of a given drug but did not account for multiple drug use. Tranquilizers lacked power for subgroup analysis.

### Antipsychotics

Both subgroups showed excellent covariate balance (e.g., ziprasidone-like: age SMD=0.00, BMI=-0.03; clozapine-like: age=-0.04, BMI=0.05).

In the ziprasidone-like subgroup, dSM and PC were increased while CAR and TAG were decreased (TAG padj=1.46E-05, dSM=3.15E-04, CAR=2.60E-03, PC=0.042); dSM was also increased under risperidone (padj=3.52E-06) and ziprasidone (padj=8.58E-06), and FA was decreased under risperidone (padj=2.86E-16) (Figure 2A; Supplementary Table S8). For the clozapine-like subgroup, six lipid classes were significantly altered (TAG padj=5.47E-24 ↑, PC=7.16E-16 ↓, dSM=2.10E-04 ↓, CAR=0.0111 ↑, PE=0.0117 ↑, LPC=0.0402 ↓); TAG and PE were increased in both drugs, LPC was decreased (mainly olanzapine), PC and dSM were decreased (clozapine), and CAR was increased (olanzapine) (Figure 2B; Supplementary Table S8).

**Figure 2.**
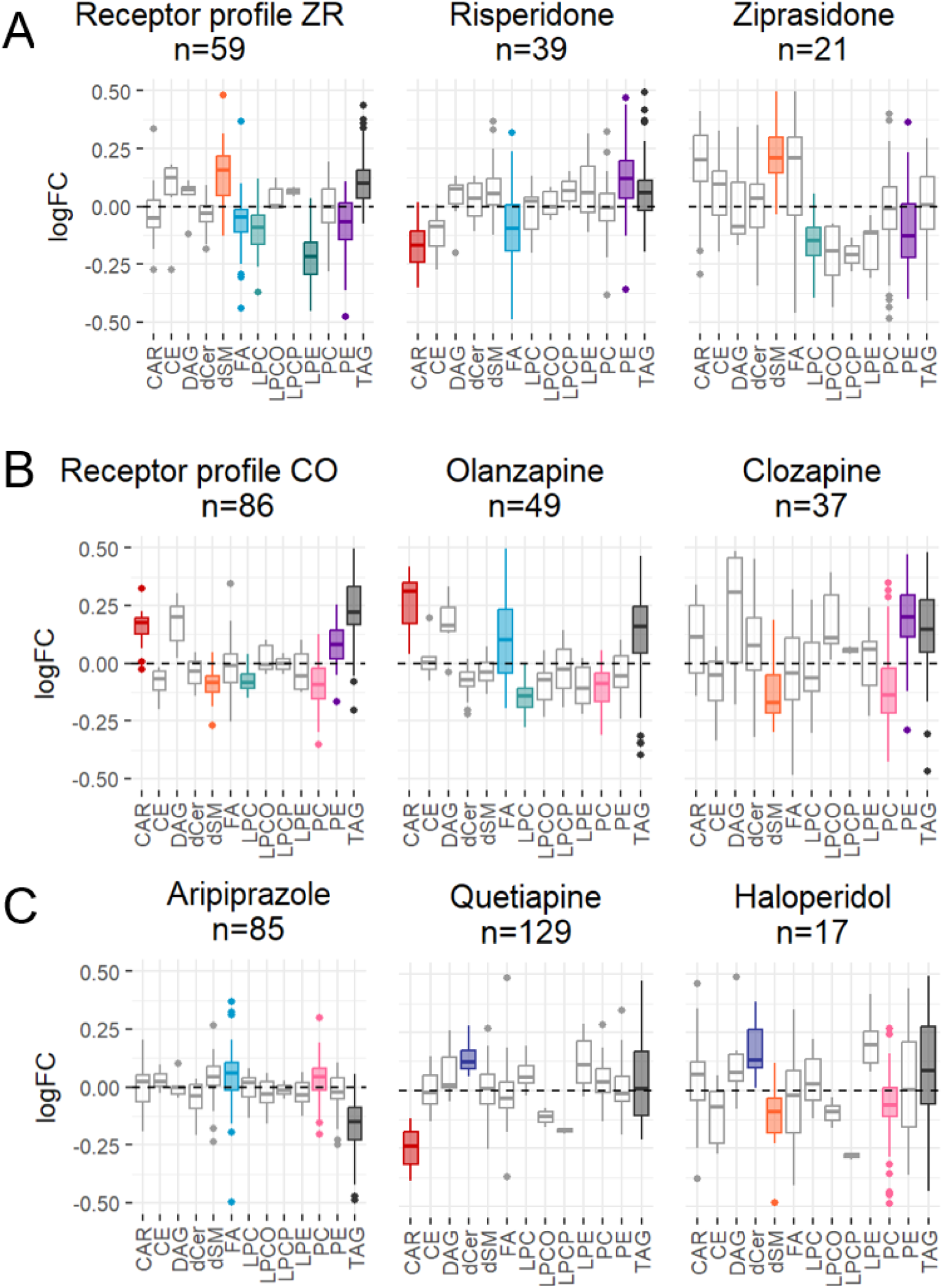
Lipid-species-level differential analysis in individuals taking antidepressants or antipsychotics Volcano plots display the results of lipid-species-based differential analysis. Each point represents one lipid species. Colored points indicate lipid species with a Benjamini-Hochberg adjusted p-value < 0.05. Antipsychotics (n = 329), Antidepressants (n = 165). Matching was performed using full matching as implemented in the MatchIt package. logFC = log fold change, CAR = acylcarnitines, CE = cholesteryl esters, DAG = diacylglycerol, dCer = ceramides, dSM = sphingomyelins, FA = fatty acids, LPC = lysophosphatidylcholines, LPCO = lysoplasmanylphosphatidylcholines, LPCP = lysophosphatidylcholine plasmalogens, LPE = lysophosphatidylethanolamines, PC = phosphatidylcholines, PE = phosphatidylethanolamines, TAG = triacylglycerides.

### Antidepressants

Covariate balance was strong for SSRIs (age SMD=0.05, BMI=0.06), good for SNRIs (sex SMD=0.19, BMI=-0.18), and fair for TCAs with BMI and diagnosis residuals; no lipid species were significant in SSRI or SNRI groups, and TCAs had n<15.

For SSRIs, PC was decreased (padj=1.02E-06), CAR increased (padj=0.0020), dSM decreased (padj=0.0077), and DAG increased (padj=0.0128); sertraline (n=35) and escitalopram (n=16) showed robust dSM decreases, opposite TAG effects (increased in escitalopram, decreased in sertraline), and individual increases in CAR, LPE, LPC, and FA (Figure 3).

**Figure 3.**
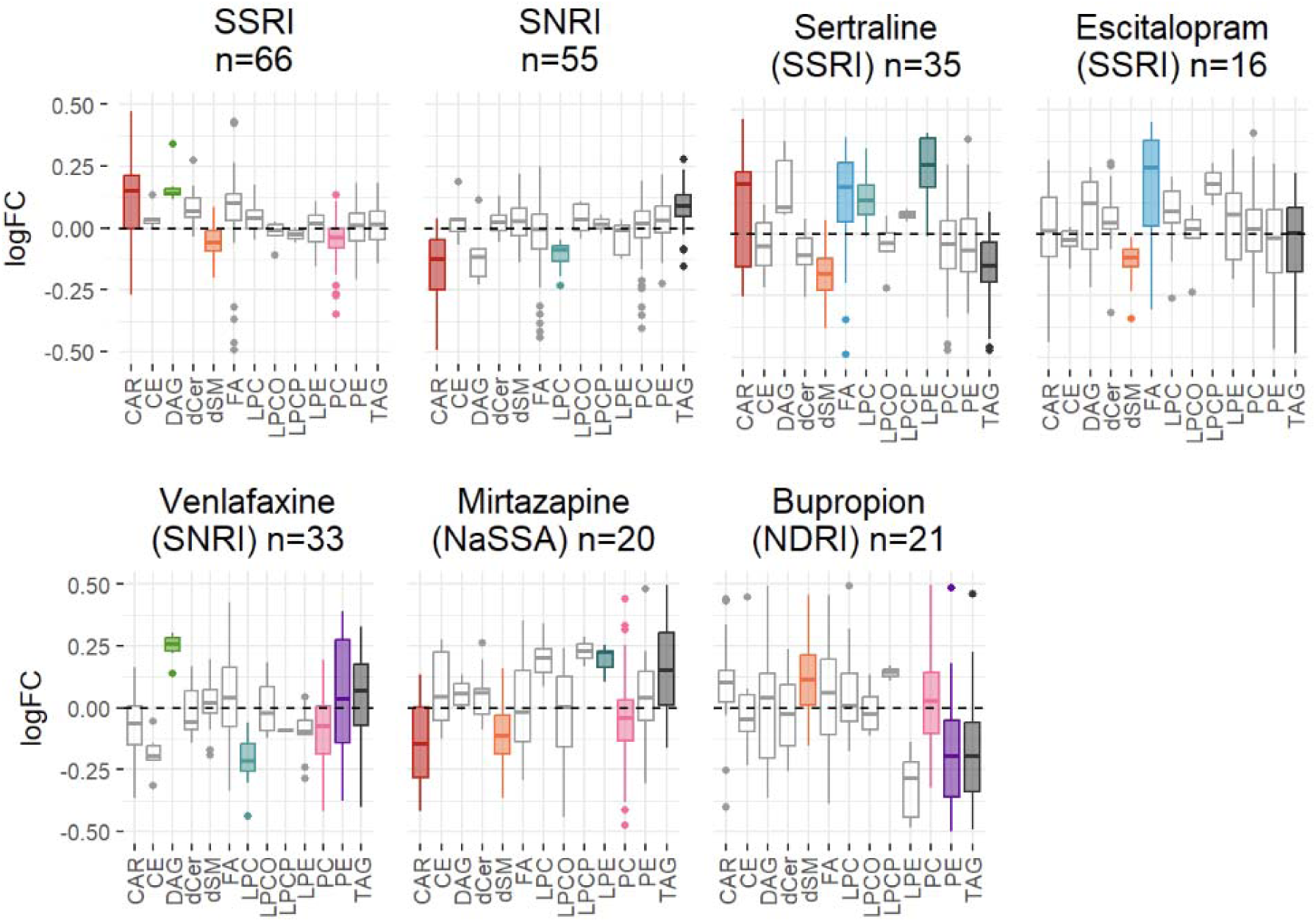
Lipid class alterations associated with individual antipsychotic treatments and the overall class Lipid class enrichment analysis in matched samples of individuals taking the overall antipsychotic class, Quetiapine, Haloperidol, Aripiprazole, Risperidone, Ziprasidone, the receptor profile CO, Olanzapine, and Clozapine. Colored boxplots represent lipid classes with significant enrichment (Benjamini–Hochberg corrected p < 0.05). Matching was performed using full matching as implemented in the MatchIt package. N = total number of individuals included in each analysis. logFC = log fold change; CAR = acylcarnitines; CE = cholesteryl esters; DAG = diacylglycerol; dCer = ceramides; dSM = sphingomyelins; FA = fatty acids; LPC = lysophosphatidylcholines; LPCO = lysoplasmanylphosphatidylcholines; LPCP = lysophosphatidylcholine plasmalogens; LPE = lysophosphatidylethanolamines; PC = phosphatidylcholines; PE = phosphatidylethanolamines; TAG = triacylglycerides.

For SNRIs, TAG was increased (padj=9.41E-09), CAR decreased (padj=7.13E-06), and LPC decreased (padj=0.0008); venlafaxine (n=33) showed increased TAG and decreased PC (Figure 3). SSRIs and SNRIs recapitulated overall antidepressant effects (TAG increased in SNRI, LPC decreased in SNRI and PC in SSRI), with additional subgroup effects on CAR, DAG, and dSM; dCer was increased overall (padj=0.0254) but power-limited in subgroups despite the same direction of effect (Figure 3; Supplementary Table S8, Figure F3).

All enrichment results for the antidepressant groups are summarized in Supplementary Table S8 and Supplementary Figure F3.

### Plasma lipidomic effects on individual drugs

Given clinical reports and our drug-class analyses showing mechanism heterogeneity does not predict lipidomic effects, individual antipsychotics and antidepressants were examined in detail.

### Antipsychotics

Seven drugs were analyzed: aripiprazole (n=162), clozapine (n=68), haloperidol (n=34), olanzapine (n=92), quetiapine (n=252), risperidone (n=76), ziprasidone (n=42). Quetiapine, olanzapine, clozapine, and aripiprazole showed broadest shifts (≥4 lipid classes significant, padj<0.05; e.g. PC, TAG, FA, CAR). TAG was significant in 6/7 drugs and PC in 4/7. Risperidone, ziprasidone, and haloperidol had more selective effects (Figure 2; Supplementary Tables S3, S8).

Consistent trends included frequent TAG increases and common FA decreases, while PC, CAR, and dSM showed heterogeneous patterns (Figure 4). Aripiprazole was unique with decreased TAG and increased FA (Figure 4).

**Figure 4.**
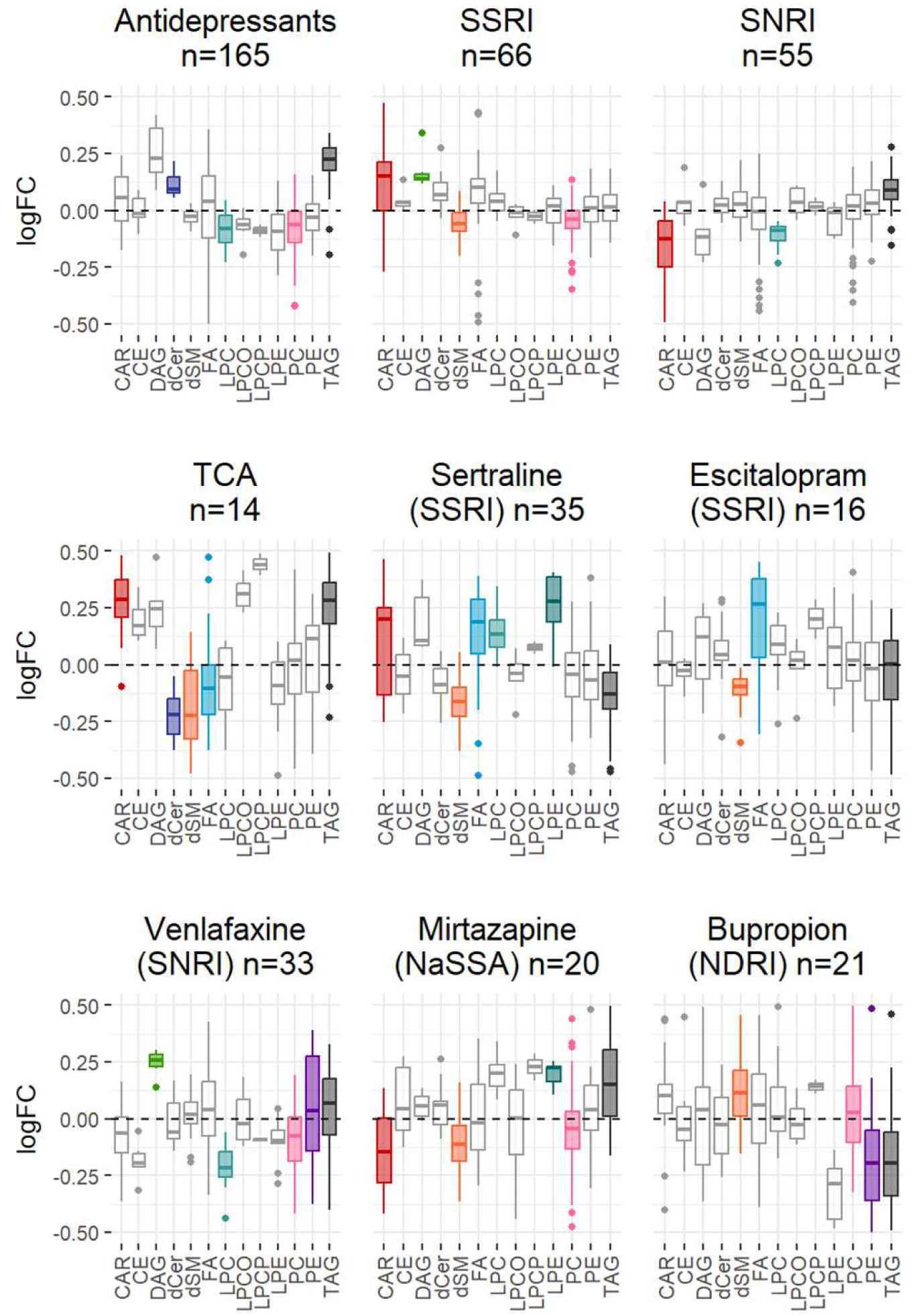
Lipid class alterations associated with individual antidepressant treatments and the overall class Lipid class enrichment analysis in matched samples of individuals taking overall antidepressant class, selective serotonin reuptake inhibitors (SSRIs), serotonin–norepinephrine reuptake inhibitors (SNRIs), tricyclic antidepressants (TCAs), Sertraline, Escitalopram, Venlafaxine, Mirtazapine, and Bupropion. Colored boxplots represent lipid classes with significant enrichment (Benjamini–Hochberg corrected p < 0.05). Matching was performed using full matdhing as implemented in the MatchIt package. N = total number of individuals included in each analysis. logFC = log fold change; CAR = acylcarnitines; CE = cholesteryl esters; DAG = diacylglycerol; dCer = ceramides; dSM = sphingomyelins; FA = fatty acids; LPC = lysophosphatidylcholines; LPCO = lysoplasmanylphosphatidylcholines; LPCP = lysophosphatidylcholine plasmalogens; LPE = lysophosphatidylethanolamines; PC = phosphatidylcholines; PE = phosphatidylethanolamines; TAG = triacylglycerides.

**Figure 5:**
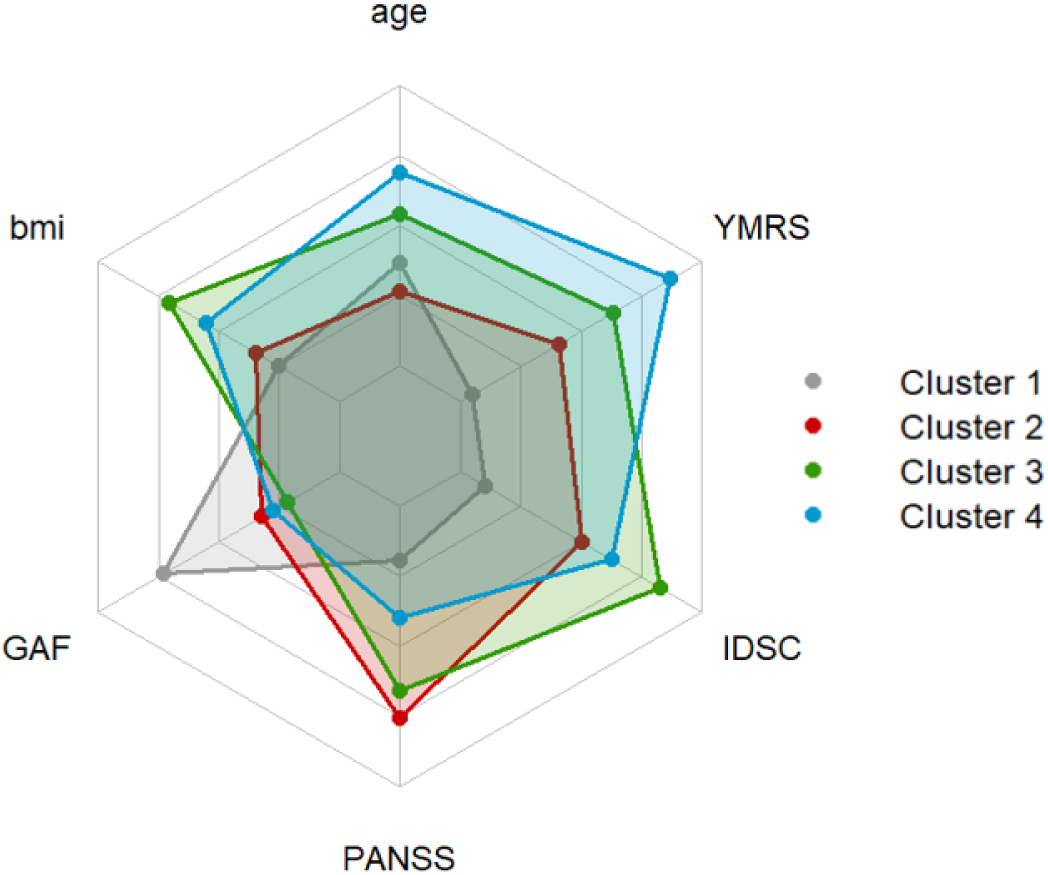
Radarplot showing the distribution of numeric covariates across the four clusters identified by hierarchical clustering: results of ANOVA analysis. BMI=body mass index, CGI=clinical global impression, GAF=global assessment of functioning, IDSC=inventory of depressive symptomatology, PANSS=positive and negative syndrome scale, YMRS=young mania rating scale.

Single-drug cohorts (patients taking only one antipsychotic, n≥15): aripiprazole (n=44), olanzapine (n=54), quetiapine (n=142), risperidone (n=30) showed no significant species but stronger, more consistent class-level effects than polypharmacy groups (e.g. clearer TAG enrichment), suggesting monotherapy reveals distinct signatures (Supplementary Table S8, Figure F4).

### Antidepressants

Five drugs were analyzed: sertraline (n=35), escitalopram (n=16), venlafaxine (n=33), bupropion (n=21), mirtazapine (n=20) (baseline characteristics: Supplementary Tables S5–S6).

Sertraline showed significant increases in TAG (padj=5.74E-09), FA (padj=3.01E-04), dSM (padj=1.71E-04), CAR (padj=3.42E-04), and LPC (padj=3.42E-04), with LPE decreased (padj=3.42E-04). TAG was significantly altered in all except venlafaxine: increased in escitalopram and mirtazapine (padj=4.26E-05, 3.07E-07), decreased in bupropion (padj=2.22E-11).

PC was increased in venlafaxine (padj=9.33E-04) and bupropion (padj=9.53E-03) but decreased in mirtazapine (padj=6.59E-03) and sertraline (padj=3.42E-04). dSM was increased in sertraline (padj=1.71E-04) and bupropion (padj=1.28E-02) but decreased in escitalopram (padj=5.97E-03) and mirtazapine (padj=6.59E-03). Venlafaxine showed increases in PC (padj=9.33E-04), LPC (padj=4.79E-03), DAG (padj=6.88E-03), and PE (padj=4.23E-03) (Supplementary Table S8, Figure F5).

Single-drug cohorts showed clearer class-level effects versus polypharmacy groups, with differences in TAG, PC and LPE directionality (venlafaxine n=33: ↑FA padj=7.73E-12, ↓TAG padj=3.93E-06, ↓LPC padj=1.54E-03; sertraline n=35: ↑CAR/TAG/FA, ↓PC padj=1.54E-09, ↓dSM; Supplementary Table S8, Figure F6).

### Medication vs. disease markers

High overlaps between SCZ and antipsychotics (92.5%), BPD and antidepressants (67.3%), and BPD and stabilizers (55.8%) confound interpretation (Tkachev et al., 2023). SCZ-associated alterations were generally stronger than antipsychotic (AP)-associated alterations. For example, TAG enrichment was stronger in SCZ (padj=9.62E-22, log2FC=0.82) than in AP users (padj=3.12E-18, log2FC=0.78), and dCer increase was more pronounced in SCZ (padj=2.10E-04, log2FC=0.77) than in AP users (padj=2.95E-03, log2FC=0.72). Nine lipid classes showed stronger enrichment in SCZ compared to four in the opposite direction (PC, CAR, DAG, PE); six of the top-10 most significantly altered lipids were more affected in SCZ (Supplementary Tables S7–S8, Figure F6; Tkachev et al., 2023).

### Hierarchical Clustering based on psychopharmacological and clinical variables

Hierarchical clustering on 593 individuals (after excluding n=29 missing covariates; mean age 41.1y, 45% female; n=177 SCZ, 228 BPD, 188 controls) used sex, age, BMI, diagnosis, and 15 drugs, yielding 4 clusters by elbow/silhouette (Supplementary Figures F1–F2; alternatives Tables S9–S10).

Clusters differed in GAF (padj=0.0061), PANSS (padj=0.0043), medication: C1 (Cluster 1; low medication: AP 23%, AD 14%, MS 23%, younger patients, mixed diagnoses); C2 (Cluster 2; moderate AP 51%, SCZ 52%, males 91%, clozapine ∼15%); C3 (Cluster 3; high AP 97%, AD 52%, high BMI); C4 (Cluster 4; BPD 96%, MS 89%, valproate 64%, older).

Lipid alterations (Supplementary Table S13): Cluster 2 showed decreased dSM (mean z=-1.25, clozapine-linked) and increased LPC/LPC_O/LPC_P (z=1.2-1.38); Cluster 3 showed increased dCer (z=0.97) and DAG (z=0.78, antidepressant-linked); Cluster 4 showed increased dSM (z=0.48); Cluster 1 showed no alterations.

ANOVA identified 206 significant lipid species. The top-5 most significant were dSMs 39:2 (padj=1.10E-17), 41:3 (2.78E-11), 32:2 (2.24E-10), plus FA 26:4 (1.80E-11) and CAR 14:0 (1.44E-10) (Supplementary Table S9). These results demonstrate that subgroups defined by diagnosis and drug treatment differ in lipid profiles. Clozapine use in Cluster 2 was clearly associated with lower dSM levels, consistent with our regression analyses and prior reports showing clozapine decreases sphingomyelin levels (Weston-Green et al., 2018). dSM and CAR classes were overrepresented (permutation p=3.00E-04, 9.00E-04). Random forest identified dSM 34:3 and 32:2 as top features (Supplementary Tables S9–S10, Figures F5/F8).

## Discussion

### Psychopharmacolipidomic alterations are both shared and unique

Our study demonstrates significant plasma lipidomic alterations in individuals treated with psychopharmacological medications compared to untreated individuals across multiple drug classes, with antipsychotics exerting the strongest and broadest effects and antidepressants producing smaller but still meaningful changes.

TAG elevations with antipsychotics align with previous reports for olanzapine, risperidone, and quetiapine (Kaddurah-Daouk et al., 2007; Almeida et al., 2020; Tkachev et al., 2023), as do changes in PCs and FAs (Kaddurah-Daouk et al., 2007; McEvoy et al., 2013). Our enrichment analysis extends these findings by highlighting less consistently reported changes in CARs and dCers, particularly with quetiapine, clozapine, and olanzapine.

Across individual antipsychotics, lipid class effects were heterogeneous: quetiapine, olanzapine, clozapine, and aripiprazole showed enrichment in four or more lipid classes, whereas risperidone, ziprasidone, and haloperidol displayed more restricted patterns. Similar differences in lipidomic effects between antipsychotics have been observed in prior clinical and experimental studies (Kaddurah-Daouk et al., 2007; Correia et al., 2021). Chen et al. reported dynamic, time-dependent lipidomic changes during quetiapine treatment in bipolar II disorder, highlighting its broad impact on plasma lipids (Chen et al., 2025). Likewise, Dai et al. identified 67 lipid species significantly altered after 8 weeks of olanzapine in first-episode schizophrenia, consistent with our findings of a strong olanzapine lipidomic signature (Dai et al., 2025). Genetic variants modulate lipid changes induced by second-generation antipsychotics (Wong et al., 2025), which may partly explain interindividual variability in lipid responses.

Monotherapy analyses confirmed TAG enrichment across most antipsychotics, but aripiprazole showed a distinct profile with reduced TAGs and increased FAs and PCs, consistent with its comparatively favorable dyslipidemia risk in clinical cohorts (Citrome et al., 2014; Zhen et al., 2016).

This divergent pattern may relate to aripiprazole’s partial D2 agonism and lower impact on hypothalamic H1 receptor expression and its selective 5-HT2C receptor agonism linked to appetite and weight gain (de Bartolomeis et al., 2015), although direct cross-study comparisons remain challenging because of methodological and exposure differences.

To our knowledge, no other studies have examined drug-related plasma lipidomic changes by specific mechanisms of action. In this study, the “ziprasidone-like” group (ziprasidone, risperidone) showed significant increases in dSM and TAG alongside decreases in FA, LPC, LPE, and PE, though changes were not uniform across all drugs. The “clozapine-like” group (clozapine, olanzapine) consistently exhibited increased TAGs, a pattern seen in nearly all antipsychotics except aripiprazole, while other lipidomic features varied.

Comprehensive human studies systematically examining plasma lipidomic changes induced by antidepressants remain limited. Nevertheless, classical antidepressants like SSRIs and amitriptyline are linked to alterations in specific lipid classes. In a rat model, long-term paroxetine and, to a lesser extent, desipramine reduced sphingosine levels in brain regions but not plasma (Jaddoa et al., 2020). The acid sphingomyelinase (Asm)-ceramide pathway in the brain mediates antidepressant effects by lowering ceramides in stress-induced depression models (Gulbins et al., 2013). Recent human studies highlight weight gain and metabolic changes linked to long-term antidepressant use, suggesting indirect lipid metabolism effects (Mouawad et al., 2025). Adjunctive ω3 PUFA supplementation in adolescents on paroxetine increased ω3 content and reduced lipid peroxidation, correlating with symptom improvement (Wang et al., 2025). Another study in healthy adults found no significant changes in clinical blood lipids with SSRIs (Shostak et al., 2025). Subgroup analyses showed SSRIs, SNRIs, and TCAs present distinct lipidomic profiles, with TAG increases most consistent and decreases in PC and dSM also observed. Although fold changes are smaller than with antipsychotics, antidepressants meaningfully modulate lipid metabolism, with sphingolipids emerging as potentially important targets. The modest changes may partly reflect disease composition, as many treated participants have BPD, where lipidomic alterations are generally less pronounced than in SCZ (Tkachev et al., 2023)

At the single-drug level, sertraline and mirtazapine increased TAGs, while venlafaxine decreased PCs and LPCs. These patterns suggest that individual antidepressants have distinct lipidomic signatures reflecting their metabolic profiles (e.g., weight gain and metabolic syndrome with mirtazapine). Across both antidepressants and antipsychotics, shared effects include TAG, PC, and dSM alterations. Certain changes correspond to drugs with similar pharmacological properties, like increased TAGs and decreased PCs in “clozapine-like” antipsychotics (clozapine, olanzapine) and sedative antidepressant mirtazapine. Unique patterns exist, including aripiprazole’s decreased TAGs with increased FAs and PCs, and divergent PC/LPC profiles in venlafaxine and bupropion, indicating plasma lipidomic effects partly independent of primary drug mechanisms.

This untargeted plasma lipidomic analysis extends known antipsychotic effects beyond clinical lipids such as dyslipidemia to reveal broad alterations in non-clinical lipid classes, suggesting dyslipidemia is the “tip of the iceberg.” These changes offer insights into drug effects and adverse reactions and may guide future drug development or repurposing targeting the lipidome in disorders like SCZ or BPD (Schulte et al., 2024)

### Pharmacologically-derived hierarchical clusters are associated with distinct lipidomic profiles

Hierarchical clustering revealed distinct lipidomic profiles linked to clinical variables (age, sex, BMI, diagnosis) and psychopharmacological drug use. Four transdiagnostic clusters differed in symptom severity (GAF, PANSS) and medication patterns, ranging from low antipsychotic/antidepressant use to high polypharmacy dominated by BPD. These clusters were associated with alterations in specific lipid species (dSM 39:2, 41:3, 32:2; FA 26:4; CAR 14:0) and lipid classes (dSM, CAR). Previous PsyCourse clustering without medication data identified subgroups with clinical and cognitive differences, including a “severe psychosis” cluster (Dwyer et al., 2021); our Cluster 3 mirrors this severe group with higher symptoms, worse function, highest antipsychotic/antidepressant use, and elevated BMI.

CARs, fatty acid metabolites and markers of mitochondrial/peroxisomal β-oxidation deficits and insulin resistance (Dambrova et al., 2022), showed pronounced reductions in Cluster 3, perhaps indicating mitochondrial dysfunction related to severe symptoms and extensive antipsychotic use including clozapine. Cluster 4, dominated by BPD and high valproate use, showed elevated CARs, possibly due to altered fatty acid metabolism. This contrasts with prior reports of CAR reductions in MDD, BPD, and SCZ (Ahmed et al., 2019; Guo et al., 2022; Nasca et al., 2018; Cao et al., 2019).

TAG levels varied by cluster, with Cluster 1 (low symptoms, BMI, minimal drug use) showing low TAGs, while Clusters 2–4 had mild TAG elevations consistent with psychosis and antipsychotic treatment links (Pillinger et al., 2017). No cluster showed uniformly high TAGs, likely due to combined clinical and medication influences obscuring distinct metabolic subgroups.

### Sphingomyelins as targets of interest

dSM are abundant sphingolipids in mammalian membranes whose saturated chains and hydrogen bonding form lipid rafts—specialized domains essential for signaling, protein sorting, and membrane trafficking (Slotte et al., 2013; Chakraborty et al., 2013). dSM also interact with cholesterol to stabilize membranes and are major components of myelin sheaths (Davis et al., 2020). Beyond structure, dSM are precursors to bioactive ceramides regulating apoptosis, proliferation, inflammation, and signaling (Slotte et al., 2013; Mühle et al., 2019).

Altered dSM metabolism is implicated in psychiatric disorders, notably SCZ and major depression (Mühle et al., 2019). Studies in rodents (Chestnykh et al., 2025) and humans (Fessel et al., 2022; Senko et al., 2024) link dSM and oligodendrocytes to SCZ pathogenesis.

Sphingomyelin synthases regulate dSM/ceramide balance, autophagy, and lipid homeostasis (Mühle et al., 2019). Antidepressants like amitriptyline and fluoxetine modulate acid sphingomyelinase activity, affecting sphingomyelin/ceramide balance and neurogenesis (Mühle et al., 2019). Clozapine reduces hepatic ceramide and sphingomyelin levels in rats, linking sphingolipid changes to therapeutic and metabolic effects (Weston-Green et al., 2018). Our study recapitulates these findings in humans, showing clozapine-associated decreases in dSM (padj = 0.0001, NES = –2.36).

dSM and related sphingolipids play roles in inflammation and metabolic comorbidities common in psychiatric patients (Mühle et al., 2019; Chakraborty et al., 2013). Their involvement in inflammatory pathways may link lipidomic alterations in SCZ and MDD to disease mechanisms and treatment response, opening avenues for treatment and drug repurposing targeting sphingolipid metabolism (Ozbayraktar et al., 2010; Guo et al., 2022; Loewith et al., 2019).

### Disentangling medication- and disease-specific effects on the lipidome

The overlap of lipidomic alterations in SCZ and antipsychotic groups suggests medication-driven changes may mask or mimic disease-specific signatures. To address this, we employed a transdiagnostic approach including all drug users regardless of diagnosis, matched sample analysis to control for diagnosis and demographics, and unsupervised clustering to identify lipidomic subgroups beyond diagnostic or drug classes.

To better understand confounding from disease-related plasma lipidomic alterations, we compared disease- and medication-related changes. We found larger fold-changes in SCZ than BPD, mirroring the predominance of SCZ in the antipsychotic group and BPD in the antidepressant group. This aligns with prior reports of SCZ-specific lipidomic alterations, including drug-naive individuals (Yan et al., 2018), which are modulated by treatment. Burghardt et al. reported distinct metabolomic profiles in SCZ patients on second-generation antipsychotics versus BPD and controls (Burghardt et al., 2015). Lipid species predictive of treatment response support that medications act on diagnosis-specific lipid backgrounds (Guo et al., 2023). Our analysis showed significant differences in several lipid species—including selected CARs (e.g., CAR 10:2), dCers, and multiple TAGs—between antipsychotic users and non-users. Importantly, some lipids, especially certain DAG and dSM species, were absent in medication-naïve patients with schizophrenia or first-episode psychosis (Tkachev et al., 2023), indicating these changes arise from antipsychotic treatment rather than disease. Conversely, lipids such as CAR 10:1, CAR 13:1, dCer 34:1, dCer 36:3, and TAG species (55:6, 56:5, 56:6) were significant in both our antipsychotic analysis and SCZ studies, suggesting overlapping disease and treatment effects.

Psychotropic medications are not uniquely linked to specific lipid classes in our data. Lipid classes like LPC, TAG, and dSM, often reported altered in SCZ and BPD, may reflect medication effects. However, some lipid species show specific associations with medication but not disease, stressing the need for species-level analysis, as treatments target particular molecular species rather than entire classes—a detail that may have been overlooked in previous studies.

PC and LPC reductions aligned with unmedicated SCZ studies (Cao et al., 2020; McEvoy et al., 2013), suggesting disease effects. TAG increases with antipsychotics, including risperidone, are consistent with prior reports (Almeida et al., 2020). dSM changes varied by drug, partially contrasting earlier findings (Almeida et al., 2020).

The lack of lipid classes uniquely linked to medication likely reflects the challenge of separating disease- and treatment-associated changes in mostly medicated, cross-sectional cohorts.

Lipid class alterations in first-episode and chronic SCZ closely resemble those seen with antipsychotic treatment. Yan et al. found increased TAG, decreased PC, and decreased CARs after antipsychotic initiation in FEP patients (Yan et al., 2018), while Wang et al. reported decreased PC and LPC with increased SM in chronic SCZ (Wang et al., 2019). Baseline reductions in CAR species are also documented (Cao et al., 2019). Our data likewise associate antipsychotic use with increased TAG, decreased PC, and decreased CAR.

At the individual drug level, quetiapine, olanzapine, and clozapine increased TAG, while aripiprazole decreased TAG, consistent with its lower metabolic risk. Most antipsychotics, including the full group, decreased PC, but aripiprazole, risperidone, and quetiapine increased PC, showing compound-specific deviations. In BPD, elevated TAGs (Ribeiro et al., 2017; Guo et al., 2022), mixed PC changes (Guo et al., 2022; Zhang et al., 2022), and increased dSM and dCer levels (Brunkhorst-Kanaan et al., 2019; Tkachev et al., 2023) have been reported. Our findings align, indicating a lipidomic signature of elevated TAG, dSM, dCer, and reduced PC in BPD, with all but dSM significant.

### Limitations

Although one of the largest datasets on drug-related plasma lipidomic effects in mental health, the naturalistic PsyCourse design poses challenges due to polypharmacy and intertwined disease- and medication-effects. While multicenter across Germany and Austria, external validation is needed, yet no comparable plasma lipidomic and medication datasets currently exist. Splitting data into "single-drug" groups resulted in small sample sizes, reducing statistical power. Matching was done for sex, age, BMI, and diagnosis, but other confounders like comorbidities, lifestyle, and genetics were not fully controlled. The cross-sectional design limits causal inference; longitudinal data would clarify temporal medication-lipid relationships. Medication dose, timing, and adherence—factors influencing metabolic effects (Sabe et al., 2023)—were not considered. Future studies should examine dose- and drug-level dependence of lipidomic changes, especially for antipsychotics.

## Conclusion

Psychopharmacologic treatments, particularly antipsychotics, cause significant changes in plasma lipids with both shared and distinct signatures across drug classes and individual medications. Antidepressants show smaller but meaningful lipidomic effects. Beyond known changes in clinical lipids (Pillinger et al., 2020; Buhagiar et al., 2019; Bozdag et al., 2024), these drugs impact fat storage and metabolism (e.g., TAGs, FAs) (Almeida et al., 2020; Kaddurah-Daouk et al., 2007; Tkachev et al., 2023), as well as cell signaling, myelination (dSMs) (Almeida et al., 2020), and cell membrane structure influencing neurotransmitter dynamics (PCs) (McEvoy et al., 2013).

Lipidomic effects overlap with pathophysiological features of SCZ and BPD, suggesting modulation of underlying disease processes. Mendelian randomization supports a causal link between lipids and BPD (Stacey D et al., Biol Psychiatry, 2024). Changes in lipid classes related to mitochondria, inflammation, and membrane integrity—such as TAG, dSM, and PC—mirror patient cohort findings. This raises the potential to repurpose lipid-modifying drugs originally developed for metabolic or inflammatory diseases to target psychiatric lipid pathways. Distinct lipidomic alterations in severely affected patients (cluster 3) highlight opportunities for novel treatments addressing both metabolic comorbidities and core neurobiological dysfunction.

## Data Availability

All data produced in the present study are available upon reasonable request to the authors

## Acknowledgements

Thomas G. Schulze is supported by the German Research Foundation (Deutsche Forschungsgemeinschaft [DFG]) within the framework of the projects www.kfo241.de and www.PsyCourse.de (SCHU 1603/4-1, 5-1, 7-1; FA241/16-1) and by the Dr. Lisa Oehler Foundation (Kassel, Germany).

Thomas G. Schulze and Peter Falkai received support via the German Center for Mental Health (BMBF 01EE2303F). Udo Dannlowski is supported by DFG (grant FOR2107 DA1151/5-1, DA1151/5-2, DA1151/9-1, DA1151/10-1, DA1151/11-1; SFB-TRR58, Projects C09 and Z02) and by the Interdisciplinary Center for Clinical Research (IZKF) of the medical faculty of Münster (grant Dan3/022/22).

Urs Heilbronner was supported by the European Union’s Horizon 2020 Research and Innovation Program (PSY-PGx, grant agreement No 945151) and DFG (project number 514201724).

Eva C. Schulte was supported by the Munich Clinician Scientist Program.

