## Supplementary Figures for "Psychopharmacolipidomics: Understanding the Global Impact of Psychopharmacologic Treatments on Plasma Lipidomic Profiles"

Annika Weber, MD^1,2^, Sergi Papiol, PhD^1,2^, Anna Tkachev, PhD^3^, Thomas Vogl, MSc^1^, Fanny Senner, MD^1,4,5^, Sabrina K. Schaupp, MSc^1^, Daniela Reich-Erkelenz, MSc^1,4^, Mojtaba Oraki Kohshour, MSc^1,6^, Farahnaz Klöhn-Saghatolislam, MD^1^, Janos L. Kalman, MD, PhD^1,4^, Urs Heilbronner, PhD^1^, Maria Heilbronner, MSc^1^, Ashley L. Comes, PhD^1^, Monika Budde, PhD^1^, Heike Anderson-Schmidt MD, PhD^7^, Kristina Adorjan, MD^1,8^, Richard Musil^4^, Jens Wiltfang, MD^7,9,10^, Carsten Spitzer, MD^11^, Max Schmauß, MD^12^, Eva Z. Reininghaus, MD^13^, Georg Juckel, MD^14^, Andreas Fallgatter, MD^15,16^, Udo Dannlowski, PhD^17^, Martin von Hagen, MD^18^, Peter Falkai, MD^4,19^, Philipp Khaitovich, PhD^3^, Thomas G. Schulze^1,19-21,#^, MD, Eva C. Schulte, MD, PhD^1,4,19,22,23,#,*^

1 Institute of Psychiatric Phenomics and Genomics (IPPG), LMU University Hospital, LMU Munich, Munich, 80336, Germany
2 Max Planck Institute of Psychiatry, Munich, 80804, Germany
3 Vladimir Zelman Center for Neurobiology and Brain Rehabilitation, Skolkovo Institute of Science and Technology, Moscow, 121205, Russia
4 Department of Psychiatry and Psychotherapy, LMU University Hospital, LMU Munich, Munich, 80336, Germany
5 Centres for Psychiatry Suedwuerttemberg, Ravensburg, 88214, Germany
6 Department of Immunology, Faculty of Medicine, Ahvaz Jundishapur University of Medical Sciences, Ahvaz, Iran
7 Department of Psychiatry and Psychotherapy, University Medical Center Göttingen, Göttingen, 37075, Germany
8 University Hospital of Psychiatry and Psychotherapy, University of Bern, Bern, Switzerland
9 German Center for Neurodegenerative Diseases (DZNE), Göttingen, 37075, Germany
10 Neurosciences and Signaling Group, Institute of Biomedicine (iBiMED), Department of Medical Sciences, University of Aveiro, Aveiro, Portugal
11 Department of Psychosomatic Medicine and Psychotherapy, University Medical Center Rostock, Rostock, 18147, Germany
12 Clinic for Psychiatry, Psychotherapy and Psychosomatics, Augsburg University, Medical Faculty, Bezirkskrankenhaus Augsburg, Augsburg, 86156, Germany
13 Division of Psychiatry and Psychotherapeutic Medicine, Research Unit for Bipolar Affective Disorder, Medical University of Graz, Graz, 8036, Austria
14 Department of Psychiatry, Ruhr University Bochum, LWL University Hospital, Bochum, 44791, Germany
15 Department of Psychiatry and Psychotherapy, Tübingen Center for Mental Health (TüCMH), University of Tübingen, Tübingen, 72076, Germany
16 German Center for Mental Health (DZPG), partner site Tübingen, Tübingen, 72076, Germany
17 Institute for Translational Psychiatry, University of Münster, Münster, 48149, Germany
18 Clinic for Psychiatry and Psychotherapy, Clinical Center Werra-Meißner, Eschwege, 37269, Germany
19 German Center for Mental Health (DZPG), partner site Munich-Augsburg
20 Department of Psychiatry and Behavioral Sciences, Norton College of Medicine, SUNY Upstate Medical University, Syracuse, NY, USA
21 Department of Psychiatry and Behavioral Sciences, Johns Hopkins University School of Medicine, Baltimore, MD, USA
22 Department of Psychiatry, University Hospital, Faculty of Medicine, University of Bonn, Bonn, Germany
23 Institute of Human Genetics, University Hospital, Faculty of Medicine, University of Bonn, Bonn, Germany

^#^	shared senior authorship

^*^	corresponding authors

Corresponding authors email:

Eva C. Schulte

##### Lipid quantification process

Non-fasting plasma samples were collected between 2012 and 2016. Liquid chromatography coupled with untargeted mass spectrometry (LC-MS) consisted of a Waters Acquity UPLC system (Waters, Manchester, UK) and a Q Exactive orbitrap mass spectrometer (Thermo Fisher Scientific, USA) equipped with a heated electro-spray ionization (HESI) probe as a custom platform at Skolkovo Technical University in Moscow, Russia. Separation of lipids was performed using a reverse phase ACQUITY UPLC BEH C8 Column (2.1 × 100 mm, 1.7 μm, Waters co., Milford, MA, USA) coupled to a Vanguard precolumn. Mass spectra were recorded in both positive and negative modes. Spectra were analyzed with the XCMS software (Smith C Ann Chem 2006), which employed the “centWave” method for peak detection (Tkachev et al)

### Supplementary Figures

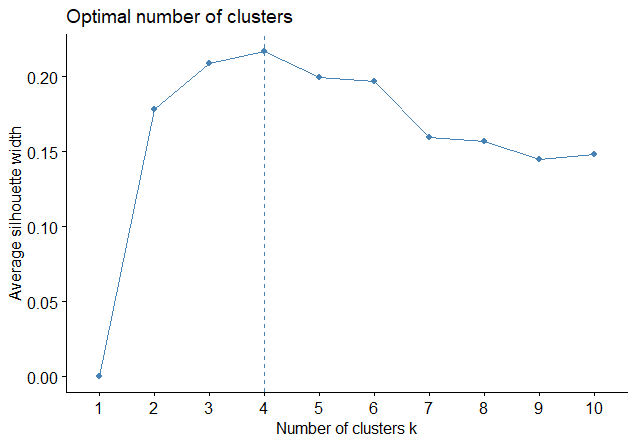

**Supplementary Figure F1**: Silhouette Plot for the full cohort.

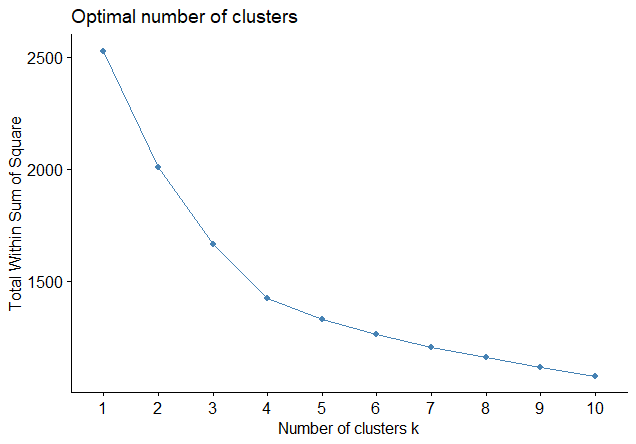

**Supplementary Figure F2**: Silhouette Plot for the full cohort.

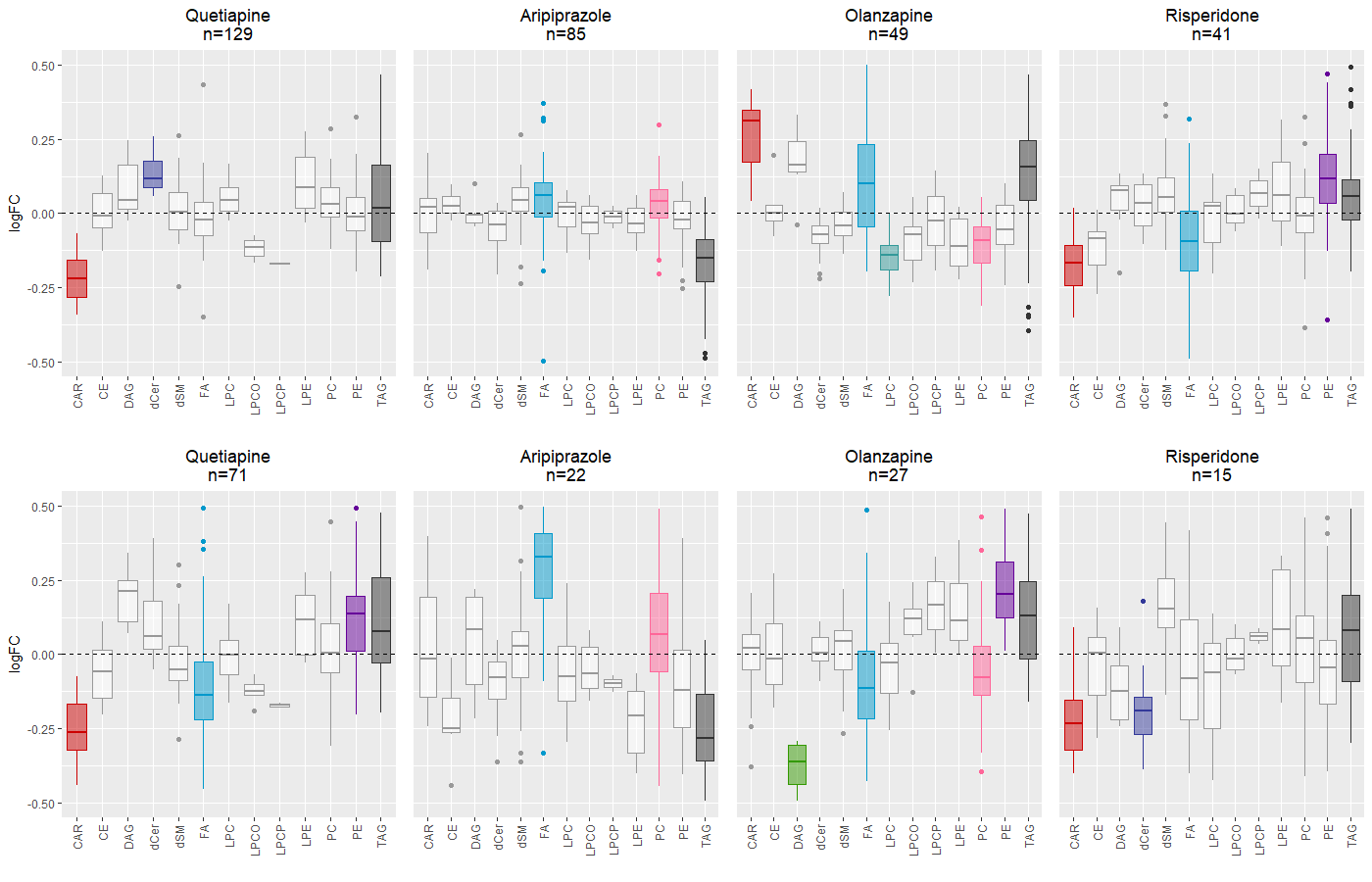

**Supplementary Figure F4**: Enrichment analysis (using LipidR) for lipid classes for single vs. multiple drug use in the matched groups for aripiprazole, olanzapine, quetiapine and risperidone.
A: all patients taking a specific drug (including those who take more than one antipsychotic drug)
B: patients taking just one antipsychotic drug
Colored boxplots represent enrichments significant at p<0.05 after Benjamini-Hochberg correction. n=total number of individuals included in the analysis. logFC=log fold change, CAR=acylcarnitines, CE=cholesteryl esters, DAG=diacylglycerol, dCer=ceramides, dSM=sphingomyelins, FA=fatty acids, LPC=lysophosphatidylcholines, LPCO= lysoplasmanylphosphatidylcholines, LPCP=lysophosphatidylcholine plasmalogens, LPE= lysophosphatidylethanolamines, PC=phosphatidylcholines, PE=phosphatidylethanolamines, TAG=triacylglycerides.

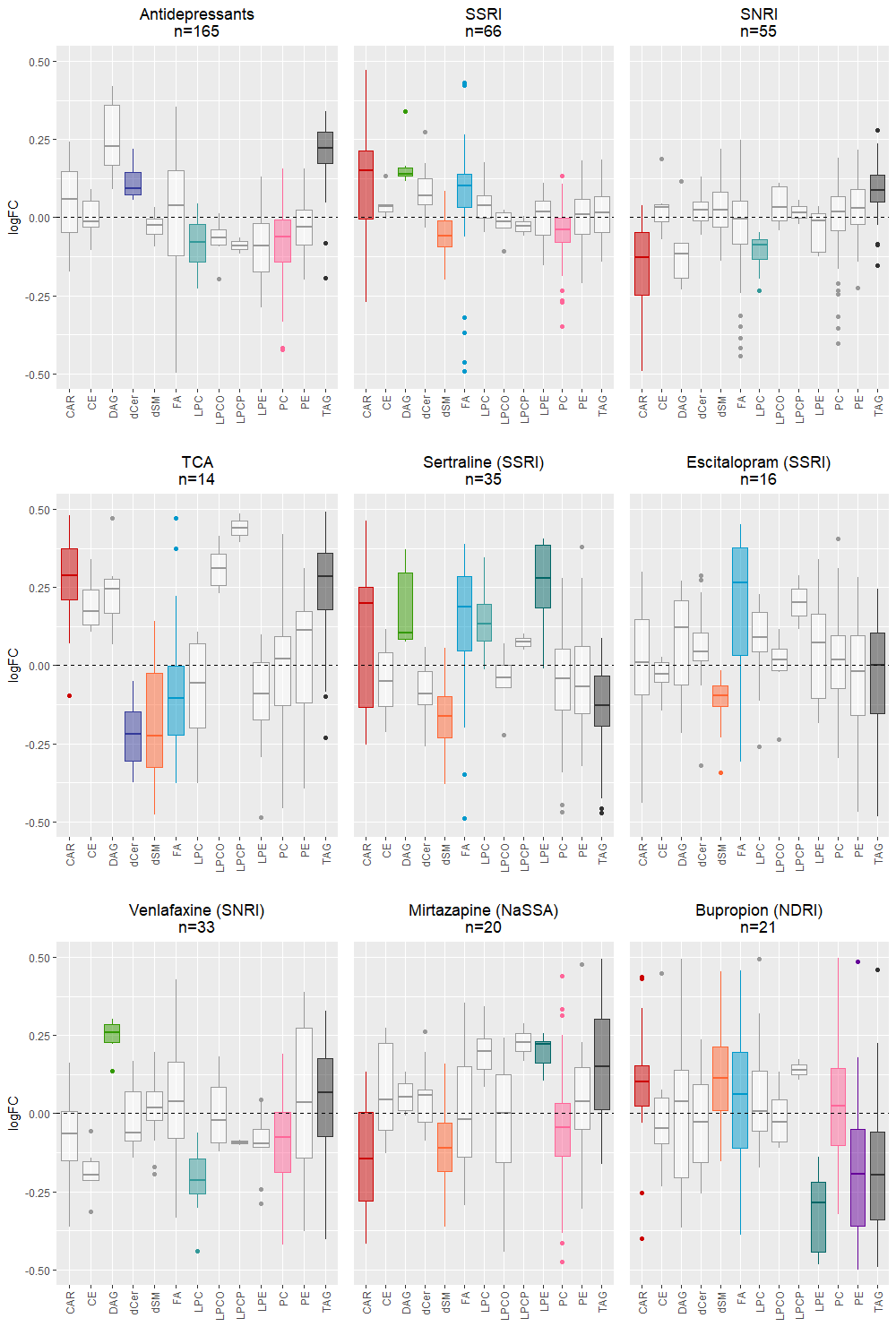

**Supplementary Figure F5:** Enrichment analysis (using LipidR) for lipid classes for antidepressants, SSRI, SNRI, TCA, sertraline, escitalopram, venlafaxine, mirtazapine and burpropion. Colored boxplots represent enrichments significant at p<0.05 after Benjamini-Hochberg correction. n=total number of individuals included in the analysis.
logFC=log fold change, CAR=acylcarnitines, CE=cholesteryl esters, DAG=diacylglycerol, dCer=ceramides, dSM=sphingomyelins, FA=fatty acids, LPC=lysophosphatidylcholines, LPCO= lysoplasmanylphosphatidylcholines, LPCP=lysophosphatidylcholine plasmalogens, LPE= lysophosphatidylethanolamines, PC=phosphatidylcholines, PE=phosphatidylethanolamines, TAG=triacylglycerides.

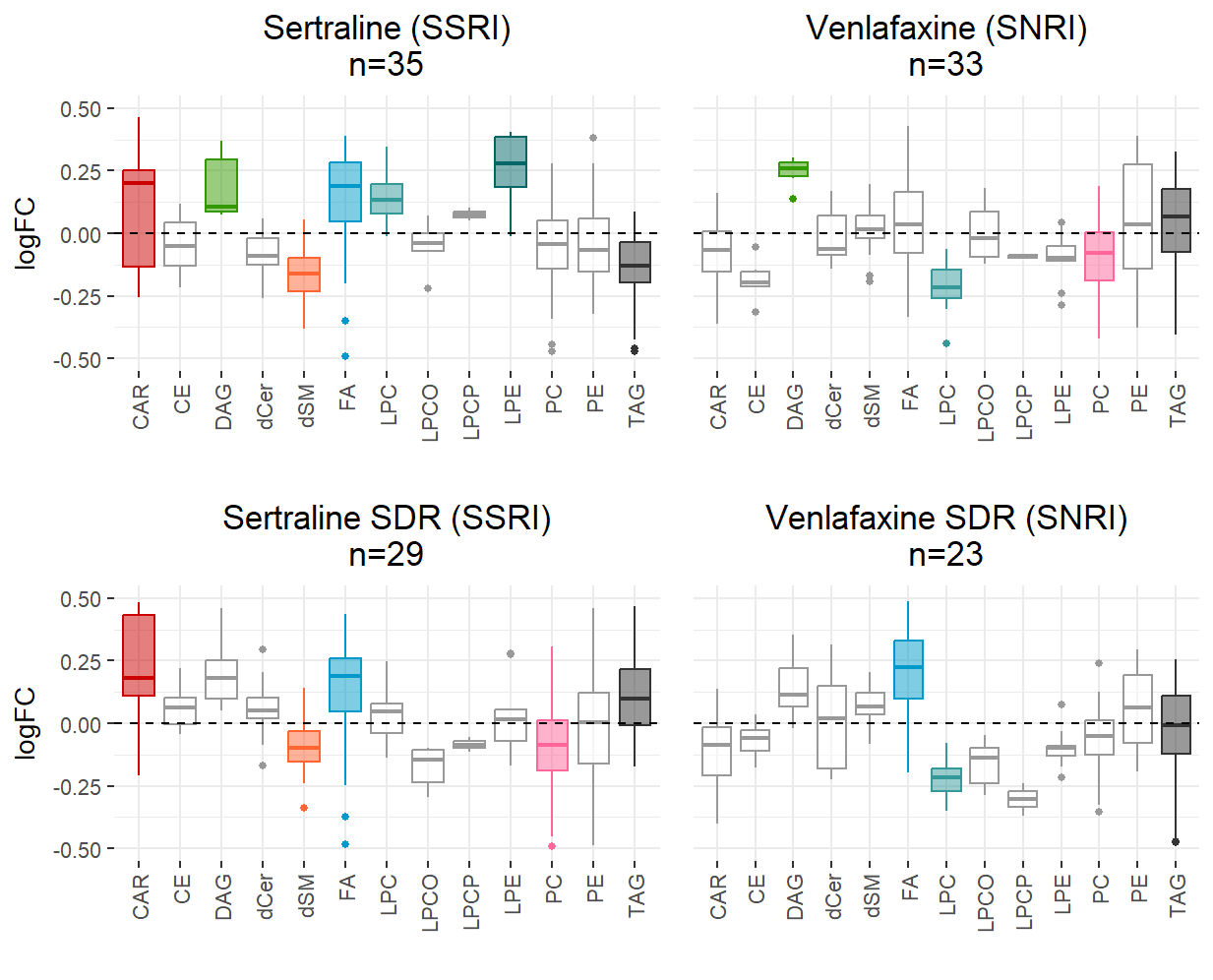

**Supplementary Figure F6:** Enrichment analysis (using LipidR) for lipid classes for sertraline and venlafaxine as well as the corresponding single drug analyses. Colored boxplots represent enrichments significant at p<0.05 after Benjamini-Hochberg correction. n=total number of individuals included in the analysis.
logFC=log fold change, CAR=acylcarnitines, CE=cholesteryl esters, DAG=diacylglycerol, dCer=ceramides, dSM=sphingomyelins, FA=fatty acids, LPC=lysophosphatidylcholines, LPCO= lysoplasmanylphosphatidylcholines, LPCP=lysophosphatidylcholine plasmalogens, LPE= lysophosphatidylethanolamines, PC=phosphatidylcholines, PE=phosphatidylethanolamines, TAG=triacylglycerides.

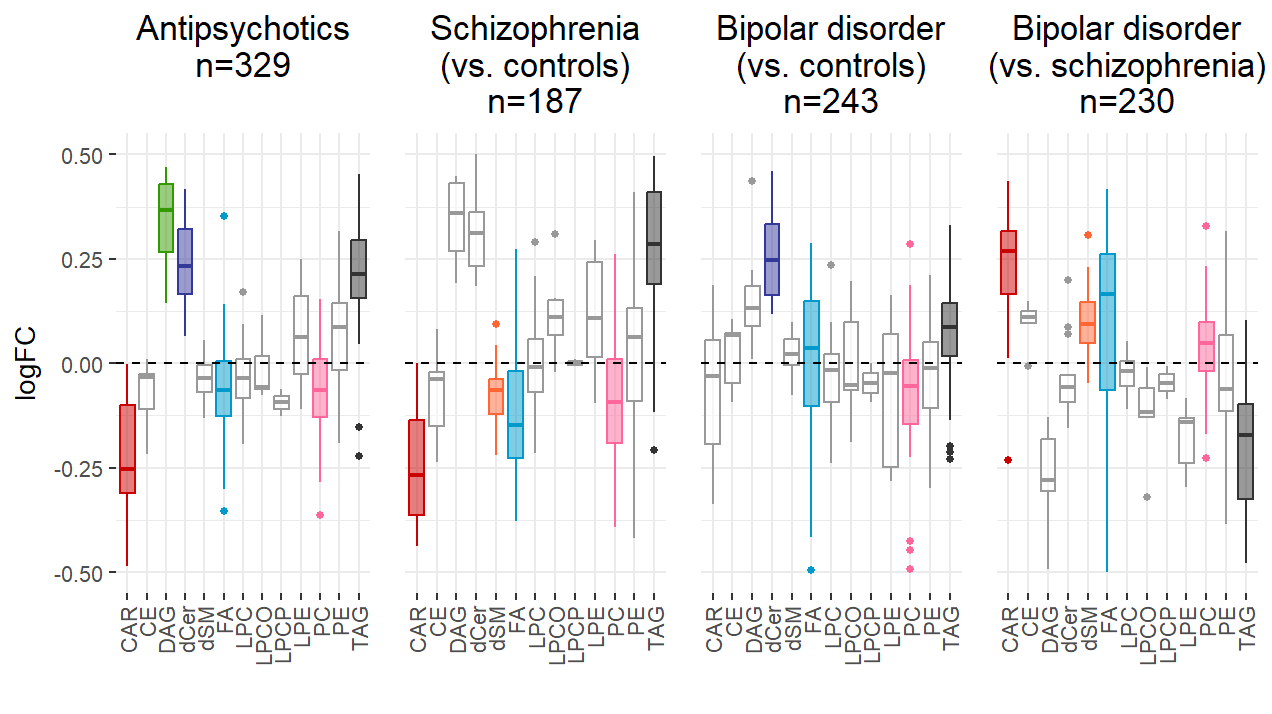

**Supplementary Figure F7:** Enrichment analysis (using LipidR) for lipid classes for AP, SCZ, BP and BP vs. SCZ. Colored boxplots represent enrichments significant at p<0.05 after Benjamini-Hochberg correction. n=total number of individuals included in the analysis. logFC=log fold change, CAR=acylcarnitines, CE=cholesteryl esters, DAG=diacylglycerol, dCer=ceramides, dSM=sphingomyelins, FA=fatty acids, LPC=lysophosphatidylcholines, LPCO= lysoplasmanylphosphatidylcholines, LPCP=lysophosphatidylcholine plasmalogens, LPE= lysophosphatidylethanolamines, PC=phosphatidylcholines, PE=phosphatidylethanolamines, TAG=triacylglycerides.

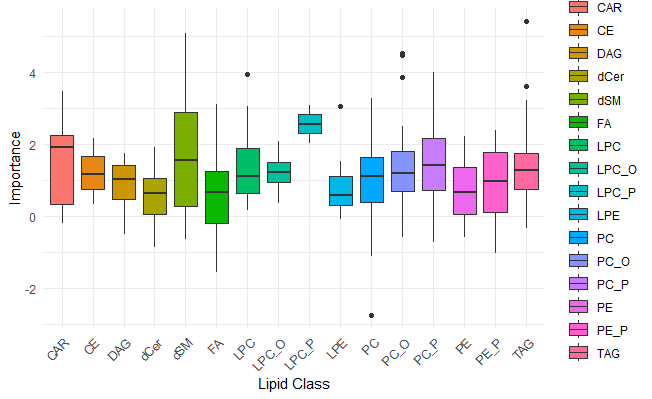

**Supplementary Figure F8:** Boxplot of lipid class importance in the random forest model for the four-cluster solution: distribution of feature importance across lipid classes. CAR=acylcarnitines, CE=cholesteryl esters, DAG=diacylglycerol, dCer=ceramides, dSM=sphingomyelins, FA=fatty acids, LPC=lysophosphatidylcholines, LPCO= lysoplasmanylphosphatidylcholines, LPCP=lysophosphatidylcholine plasmalogens, LPE= lysophosphatidylethanolamines, PC=phosphatidylcholines, PE=phosphatidylethanolamines, TAG=triacylglycerides
